# Deregulated cellular circuits driving immunoglobulins and complement consumption associate with the severity of COVID-19

**DOI:** 10.1101/2020.06.15.20131706

**Authors:** Ana Marcos-Jiménez, Santiago Sánchez-Alonso, Ana Alcaraz-Serna, Laura Esparcia, Celia López-Sanz, Miguel Sampedro-Núñez, Tamara Mateu-Albero, Ildefonso Sánchez-Cerrillo, Pedro Martínez-Fleta, Ligia Gabrie, Luciana del Campo Guerola, Margarita López-Trascasa, Enrique Martín-Gayo, María Calzada, Santos Castañeda, Hortensia de la Fuente, Isidoro González-Álvaro, Francisco Sánchez-Madrid, Cecilia Muñoz-Calleja, Arantzazu Alfranca

## Abstract

**Background:** SARS-CoV-2 infection causes an abrupt response by the host immune system, which is largely responsible for the pathogenesis and outcome of COVID-19. We aimed to investigate which specific responses from either cellular or humoral immunity associate to severity and progression of COVID-19.

**Methods:** A cohort of 276 patients classified in mild, moderate and severe, was studied. Peripheral blood lymphocyte subpopulations were quantified by flow cytometry, and immunoglobulins and complement proteins by nephelometry.

**Results:** At admission, dramatic lymphopenia of T, B and NK cells associated to severity. However, only the proportion of B cells increased, while T and NK cells appeared unaffected. Accordingly, the number of plasma cells and circulating follicular helper T cells (cTfh) increased, but levels of IgM, IgA and IgG were unaffected. When degrees of severity were considered, IgG was lower in severe patients, suggesting an IgG consumption by complement activation or antibody-dependent cellular cytotoxicity (ADCC). Activated CD56-CD16+ NK-cells, which mediate ADCC, were increased. Regarding complement, C3 and C4 protein levels were higher in mild and moderate, but not in severe patients, compared to healthy donors. Moreover, IgG and C4 decreased from day 0 to day 10 in patients who were hospitalized for more than two weeks, but not in patients who were discharged earlier.

**Conclusion:** Our study provides important clues to understand the immune response observed in COVID-19 patients, which is probably related to viral clearance, but also underlies its pathogenesis and severity. This study associates for the first time COVID-19 severity with an imbalanced humoral immune response characterized by excessive consumption of IgG and C4, identifying new targets for therapeutic intervention.

## Introduction

Novel coronavirus disease (COVID-19), due to severe acute respiratory coronavirus 2 (SARS-CoV-2), is either asymptomatic or presents with mild symptoms in a majority of individuals. However, up to 20% patients develop a severe form of the disease with pneumonia, which in some cases results in acute respiratory distress syndrome (ARDS) and requires invasive mechanical ventilation. ARDS, together with myocardial damage, are main causes of mortality in COVID-19(1).

A pathogenic hallmark of ARDS is the disruption of the alveolar-capillary barrier and a subsequent increase in permeability, which has been partially attributed to a maladaptive immune response. Thus, lung alveolar macrophages may be infected by the virus and become activated, leading to a cytokine release syndrome, which contributes to endothelial injury with the recruitment and activation of innate and adaptive immune cells and the extravasation of plasma components of the humoral immunity(2).

Humoral immunity against viruses plays a key role in the control of initial infection and cell-to-cell spread. It is mediated by the complement system and the immunoglobulins (Ig)(3, 4). Antibodies of the IgM isotype efficiently activate the classical cascade of complement, which is one of the most effective arms of the host defense against viruses. Secreted IgA antibodies neutralize viruses within the mucosa of the respiratory and gastrointestinal tracts. Finally, antibodies of the IgG isotype opsonize viral particles and mediate their clearance by phagocytes or antibody-dependent cell cytotoxicity (ADCC). Isotype switching and affinity maturation of antibodies, as well as the generation of memory B cells and long-lived plasma cells, are B cell responses driven by helper T cells, in particular follicular helper T cells (Tfh).

Different SARS-CoV-2 components such as pathogen-associated molecular patterns (PAMPs) and N protein, together with C reactive protein (CRP) from plasma, may activate either the alternative or the lectin pathways of the complement cascade early during infection(5). In a more advanced stage of the disease, specific anti-viral Ig and immune complexes may trigger the classical pathway of complement system, or bind Fc receptors on NK and phagocytes. NK cells are major innate immunity mediators during the anti-viral response, since they kill infected cells through different mechanisms, including ADCC, mediated by FcγRIIIA (CD16) binding to clustered IgG displayed on cell surface of virally infected cells(6).

However, deregulated humoral immune response can damage host tissues. Complement-mediated tissue injury is elicited by an intense inflammatory loop secondary to C3a- and C5a-mediated recruitment and activation of neutrophils, monocytes, macrophages, lymphocytes, and platelets. Phagocytes in turn generate reactive oxygen species (ROS) and proteases, and neutrophils release neutrophil extracellular traps (NETs), which exacerbate tissue damage(7-10).

Several studies on SARS-CoV2 infection have attempted to elucidate phenotypic features of immune cell subsets either associated with severity or predictive of disease outcome. However, these studies have been conducted, in most cases, with a limited number of patients, and clinical parameters and disease severity are not homogeneously recorded in all of them. Likewise, although specific antibodies and complement activation have been proposed to mediate some of the most severe complications of coronavirus infections, including that of SARS-CoV-2(2, 5, 11, 12), solid evidence on the role of humoral immunity effector mechanisms in the pathogenesis of SARS-CoV-2-associated ARDS is clearly needed.

## Results

### COVID-19 patients have increased B cells and plasma cells in peripheral blood

Our cohort of COVID-19 patients included 276 SARS-CoV-2 infected individuals who were further classified according to the severity of their clinical signs and symptoms in mild, moderate and severe, following recently described criteria(13). Table 1 shows their main demographic and laboratory characteristics. The median (percentile 25 and 75) age was 63 (53.25-75), 163 (59.05%) were men. The mean duration of symptoms before admission was 7.36 ± 5.2 days.

**Table I.**
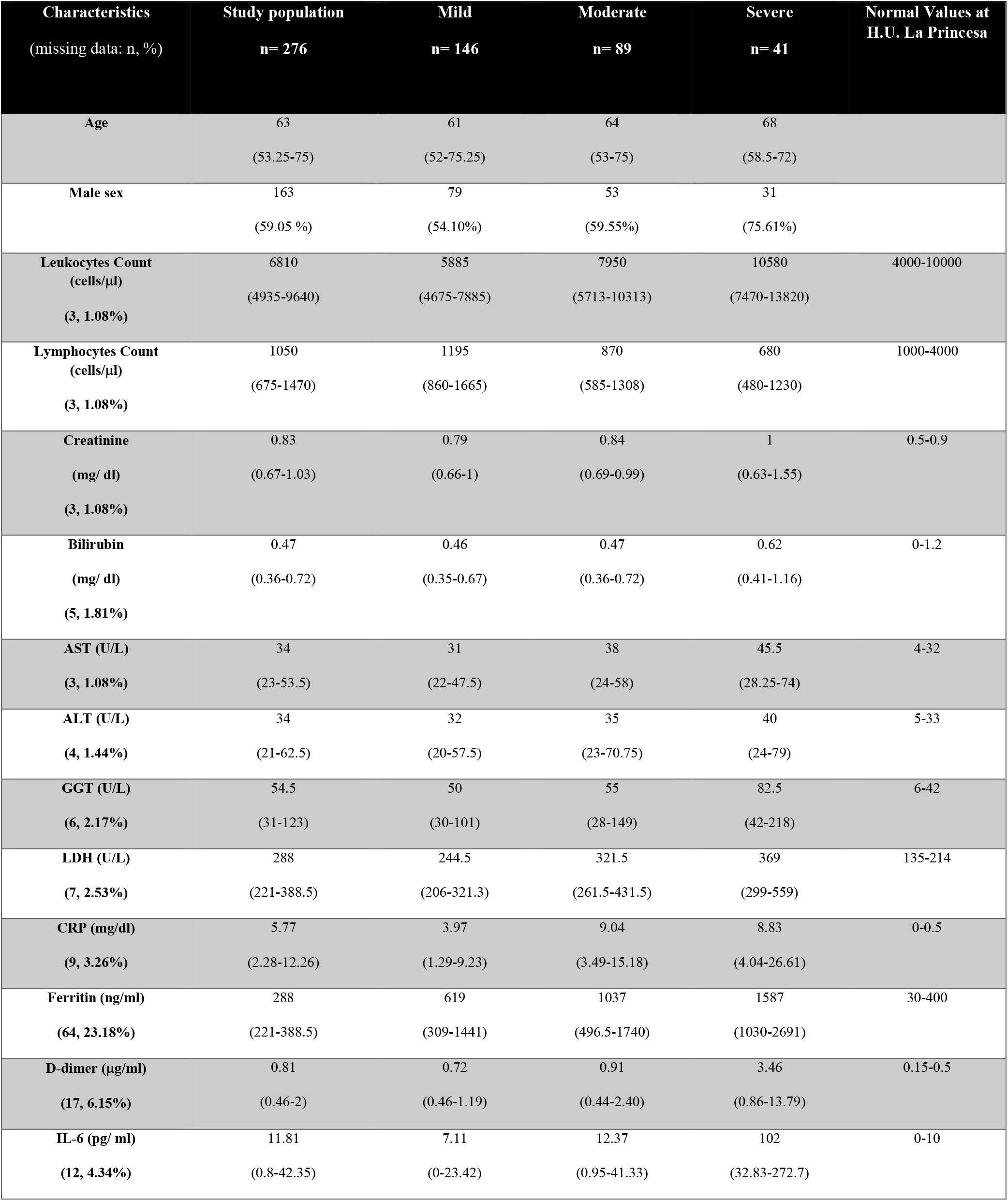
Demographic and laboratory characteristics of the study population classified by severity degree. All variables are expressed as median (p25-p75). AST: aspartate amino-transferase; ALT: alanine amino-transferase; GGT: gamma-glutamyl transferase; LDH: lactate dehydrogenase; CRP: C-reactive protein; IL-6: interleukin-6.

We conducted an initial analysis following the COVID-19 admission protocol, which comprised the quantification of the main peripheral blood lymphocyte subsets, including T, B and NK lymphocytes as well as plasma cells, by multi-parametric flow cytometry (Fig.1A, Suppl. Fig 2). The proportion of T lymphocytes (either CD4+ or CD8+) and NK cells was similar in all the patients and healthy volunteers, with the exception of CD8+ T cells, which appeared decreased in severe patients (Fig.1B). Conversely, the percentage of B cells was higher in COVID-19 patients and increased with disease severity, raising from a mean of 9.15% in healthy donors to 20.49% in severely ill patients. In accordance, plasma cells were remarkably higher in patients, both in relative and absolute values (mean 1.51% vs 17.98%; and 2.83 cells/ul vs 27.37 cells/ul, respectively). This increase in absolute plasma cell number is of special relevance, given the lymphopenia present in COVID-19 patients (Table 1), and the diminished absolute number of other lymphocyte subsets (Fig.1B). The elevated number of plasma cells suggested a redistribution of the main maturation stages of the B lineage, including naïve, transitional, unswitched memory, IgM-only memory and class-switched memory B-cells, which were identified in a subgroup of 84 patients from our initial cohort, with the gating strategy shown in Suppl. Fig 1. In COVID-19 patients, the proportion of IgM-only memory B-cells increased, while unswitched memory cells decreased (Fig.1C). This redistribution was more marked in mild patients and progressively diminished in moderate and severe patients. Nevertheless, differences among patients with different severity degree were not statistically significant (Suppl. Fig 3). No other differences in absolute numbers or percentages in other B-cell subsets could be observed between healthy donors and COVID-19 patients.

**Figure 1.**
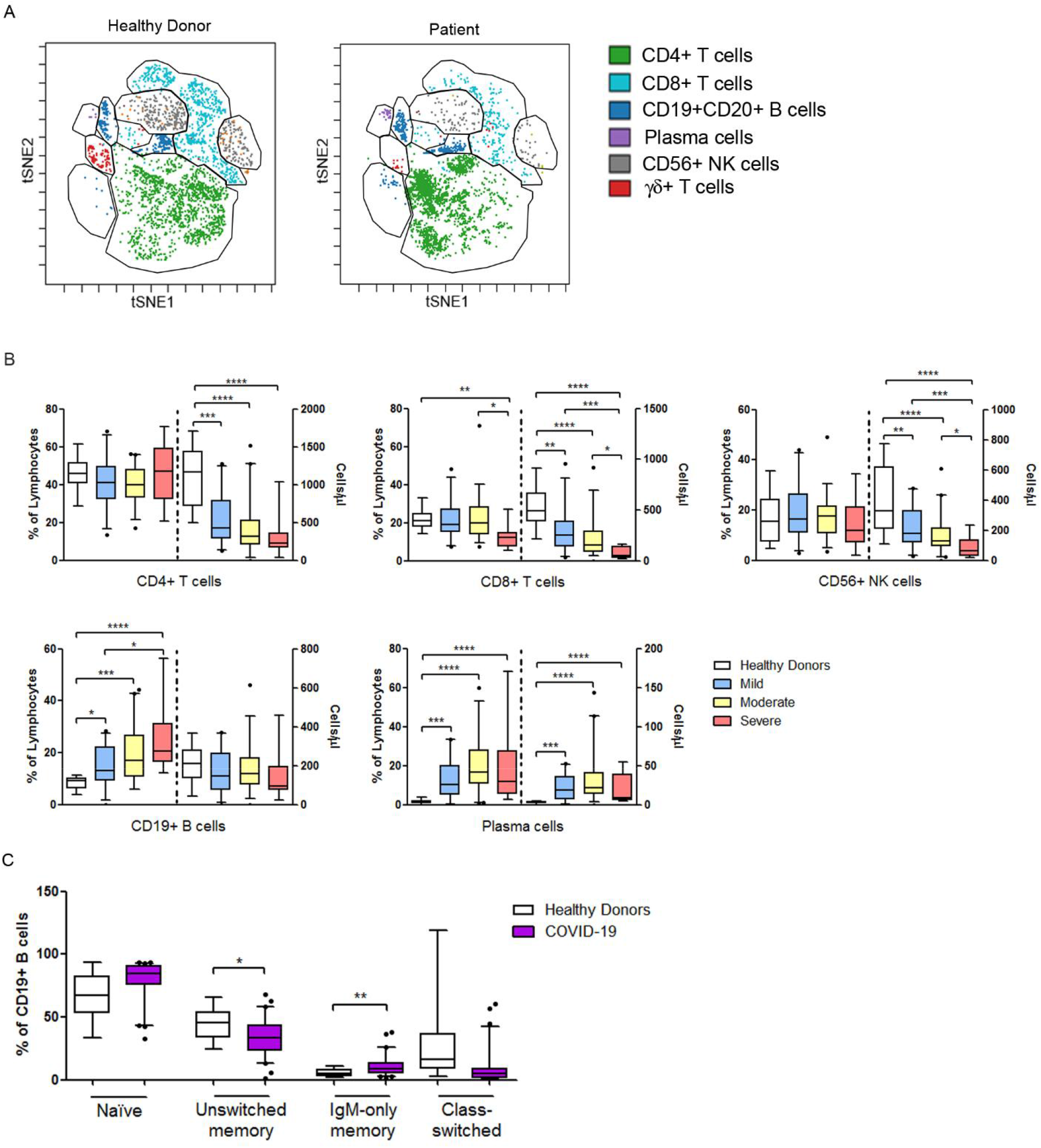
Distribution of lymphocyte subpopulations in the peripheral blood of COVID-19 patients characterized by multiparametric flow cytometry. **(A)** Analysis of distribution of lymphocyte subpopulations by automated clustering and dimensionality reduction FlowSOM tool (Cytobank) in a representative healthy donor and COVID-19 patient. **(B)** Boxplots show percentage and absolute number (cells/ul) of distinct lymphocyte subpopulations in healthy donors (white; n=19) and COVID-19 patients with different degrees of severity: mild (blue; n= 146), moderate (yellow; n= 89) and severe (red; n= 41). **(C)** Boxplots represent quantification of relevant B cell subpopulations in healthy donors (n= 19) and selected COVID-19 patients (n= 84). Asterisks indicate significant differences (p-values for ANOVA Tukey’s contrast test: *p < 0.05, **p < 0.01, ***p < 0.001, **** p < 0.0001).

**Figure 2.**
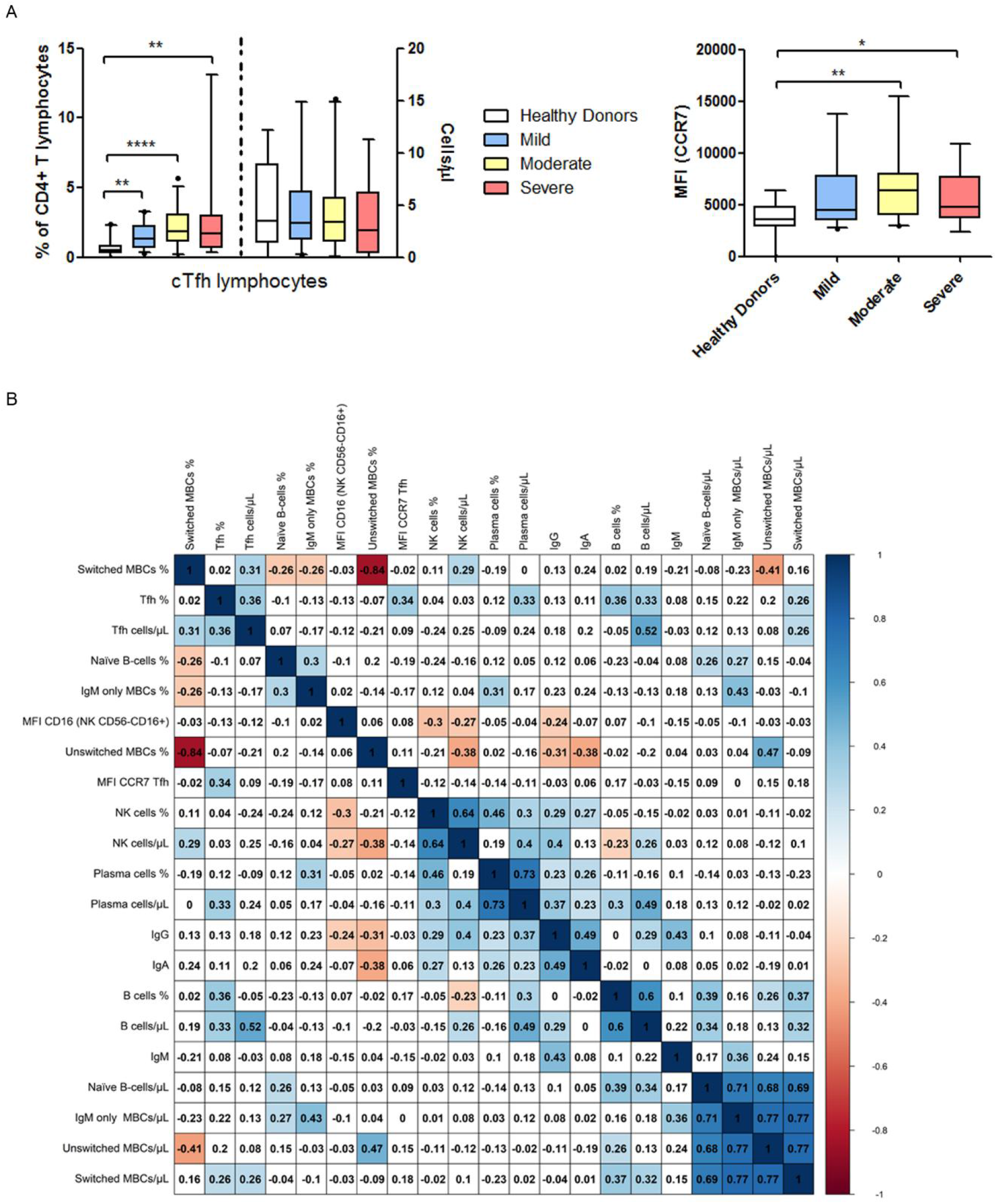
Circulating Tfh lymphocytes are increased in COVID-19 patients. **(A)** Left: Boxplots show quantification by multiparametric flow cytometry of cTfh lymphocytes as percentage or absolute number (cells/ul) in healthy donors (n= 19) and selected COVID-19 patients with different severity degree (mild (n= 29), moderate (n= 40) and severe (n= 15)). Right: Boxplots show CCR7 MFI of cTfh lymphocytes in healthy donors and COVID-19 patients with different severity degree (mild, moderate and severe). Asterisks indicate significant differences (p-values for ANOVA Tukey’s contrast test: *p < 0.05, **p < 0.01, ***p < 0.001, **** p < 0.0001) **(B)** Annotated heatmap of a correlation matrix for different variables in selected COVID-19 patients (n= 84). White squares include non-significant correlations (p > 0.05), red and blue squares include significant indirect and direct correlations (p < 0.05), respectively. Numbers inside squares and intensity of color correspond to Spearman’s rank correlation coefficient. Variables in the correlation map were reordered using hierarchical cluster method.

**Figure 3.**
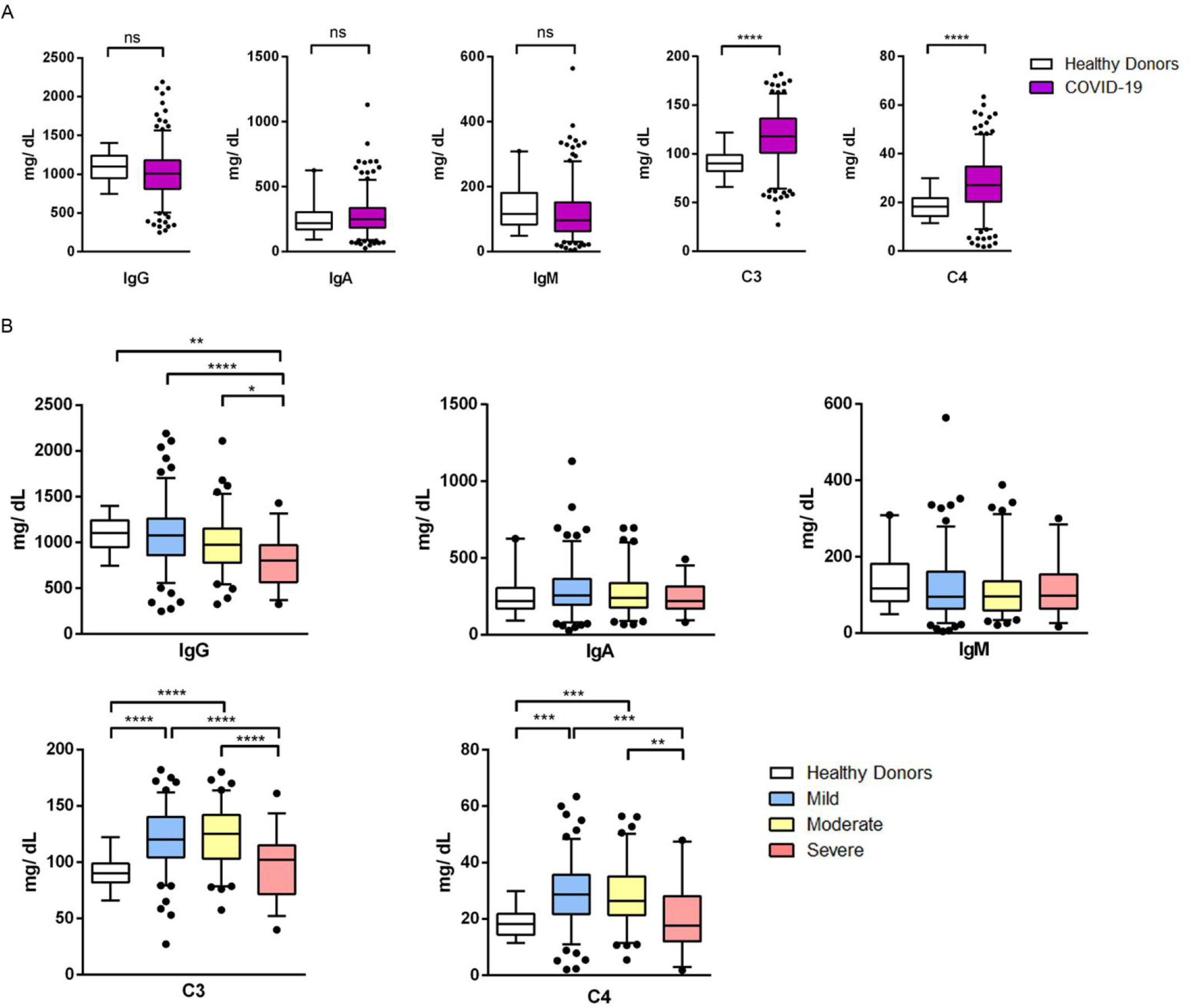
Immunoglobulins and complement levels are altered in COVID-19 patients. **(A)** Quantification of serum concentration (mg/dL) by nephelometry of IgG, IgA, IgM, C3 and C4 in healthy donors (n= 19) and COVID-19 patients (n= 37). **(B)** Serum concentration (mg/dL) of IgG, IgA, IgM, C3 and C4 in healthy donors (n=19) and COVID-19 patients according to severity degree (mild (n= 138), moderate (n= 82) and severe (n= 35). Values represent quantification for each serum marker depicted as boxplots. Asterisks indicate significant differences (p-values for Mann-Whitney t-test or ANOVA Kruskal-Wallis test, as appropriate: *p < 0.05, **p < 0.01, ***p < 0.001, **** p < 0.0001).

### Increased T follicular helper cells in the peripheral blood of COVID-19 patients

Given the role of Tfh in maturation and activation of B cells, we tested whether circulating (cTfh) were increased in the peripheral blood of COVID-19 patients, in accordance with the increased number of plasma cells. We observed that the cTfh proportion significantly increased with the severity of COVID-19 individuals, shifting from a median of 0.51 in healthy donors to 1.7 % in severely ill patients. Importantly, the absolute number remained steady despite the profound decrease of total CD4+ T cells (Fig.2A, Fig.1B). To further characterize this population, we assessed surface expression of CCR7 chemokine receptor, which has been related to lower B-cell activation capacity by Tfh(14). We found a significant increase of CCR7 expression in cTfh cells of patients with moderate to severe disease (Fig 2A). Finally, we found a direct correlation between cTfh proportion and total B-cells (r 0.33; p= 0.0090), class-switched B-cells (r 0.26, p= 0.0433), and plasma cells (r 0.33, p= 0.0091) in peripheral blood (Fig.2B).

### COVID-19 severity associates to serum levels of immunoglobulins and complement

The high number of plasma cells prompted us to investigate possible alterations in Ig concentrations. At the time of admission, COVID-19 patients had serum concentrations of either IgG, IgA or IgM isotypes comparable to those in healthy volunteers (Fig.3A). On the other hand, when patients with different degree of severity were analysed, we observed that, in severe cases, the levels of IgA and IgM were similar, but the IgG concentration was decreased, compared to healthy volunteers (Fig.3B). In addition, a direct correlation was found between IgG (0.37; p=0.0007) and IgA (0.23; p=0.0444) serum concentrations and the absolute number of plasma cells in peripheral blood, and between IgM levels and total number of peripheral blood IgM-only memory B-cells (0.36; p=0.0040) in COVID-19 patients. On the contrary, IgG (−0.31; p=0.0147) and IgA (−0.38; p=0.0021) were inversely related to the proportion of unswitched memory B cells (Fig.2B).

The increase in plasma concentrations of IL-6 and acute phase reactants that characterizes COVID-19 (Table 1) suggested that the concentration of C3 and C4 complement proteins, considered as acute phase reactants, could also be elevated. Therefore, we measured C3 and C4 levels in the sera of these patients, and could detect increased levels of both complement proteins (Fig.3A). However, when considering different groups of severity, we observed that C3 and C4 levels increased in patients with mild to moderate disease, while returned to levels similar to healthy donors in those with severe disease (Fig.3B). Therefore, in contrast to other inflammatory parameters such as LDH, ferritin or CRP (Table 1), C3 and C4 values decreased as the severity increased.

Next, we tested whether the decrease in these components of humoral immunity was actually related to the severity of the disease in critical patients, or rather mirrored an increased consumption along time. To investigate this possibility, we collected the blood of a subgroup of 37 COVID-19 patients who were still hospitalized 10 days after admission and compared it to the initial blood test. We then considered a hospitalization period longer than 15 days as a readout of the severity of the disease. With this approach, we observed that antibodies of the IgG, IgA and IgM isotypes as well as C3 and C4 complement proteins were similar in all patients at the time of admission (Fig.4A). However, after 10 days, the concentration of IgG in serum was significantly lower in patients hospitalized longer than 15 days, whereas levels of IgA and IgM did not change over this time (Fig.4A). Similarly, C3 serum levels were stable over time in both groups of patients. On the other hand, a significant decrease of C4 concentration was observed after 10 days specifically in those patients whose severity eventually required a longer stay (Fig.4A). Likewise, plasma cells, but not B lymphocytes, were significantly reduced in patients with prolonged hospitalization periods (Fig.4B).

**Figure 4.**
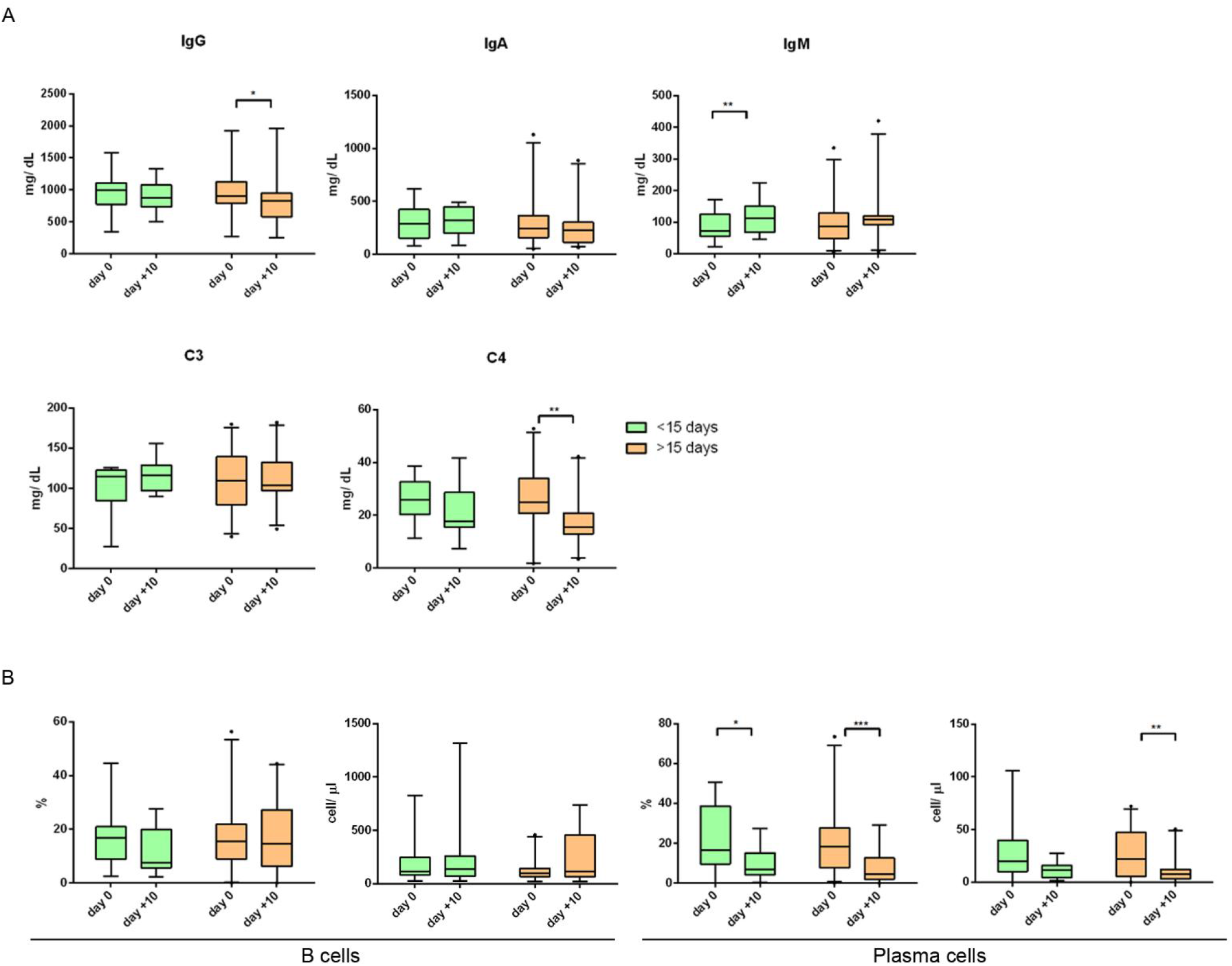
Humoral response is related to severity in COVID-19 patients. **(A)** Serum concentration (mg/dL) of IgG, IgA, IgM, C3 and C4 in COVID-19 patients according to their hospitalization period was quantified by nephelometry. **(B)** Quantification of B cells (left) and plasma cells (right) in COVID-19 patients, as percentage or absolute numbers (cells/µl), according to their hospitalization period. In both panels, X-axis indicate days from admission. Green boxplots correspond to patients with a hospitalization period of less than 15 days (n= 13) and orange boxplots to those hospitalized more than 15 days (n= 24). Asterisks indicate significant differences (p-values for Mann-Whitney t-test or Wilcoxon t-test, as appropriate: *p < 0.05, **p < 0.01, ***p < 0.001, **** p < 0.0001).

The decrease in C4 and IgG serum concentrations in severe COVID-19 cases led us to hypothesize that antigen-antibody complexes were forming in excessive amounts and activating the classical pathway of complement system. Therefore, we quantified immune complexes by enzyme immunoassay in the serum of the SARS-Cov-2 infected individuals with different levels of severity, but we did not find detectable levels of immune complexes (data not shown).

### Phenotypic features of NK cells from severe COVID-19 patients

The absence of detectable changes in circulating immune complexes led us to investigate whether antibodies of the IgG isotype could be indicative of ADCC responses. Therefore, we studied the activation profiles of NK cells in COVID-19 patients. To this end, a specific panel of antibodies was designed to quantify the main functional subsets of NK cells (CD56^bright^CD16-, CD56^dim^CD16+, CD56-CD16+). Phenotypic analysis showed a two-fold increase in the proportion of CD56-CD16+ NK cells in severe COVID-19 samples compared to healthy donors and patients with mild disease (Fig. 5A). This NK subset specializes in ADCC(15), and in accordance to its activation status, the cells showed a significant downregulation of CD16 expression irrespective of the degree of severity (Fig.5B). Interestingly, CD16 level in CD56-CD16+ cells inversely correlated to the serum concentration of IgG in COVID-19 patients (−0.24; p=0.0496) (Fig.2B). Finally, a tendency towards an increased expression of the cytotoxic marker CD107 was observed in this population (data not shown).

**Figure 5.**
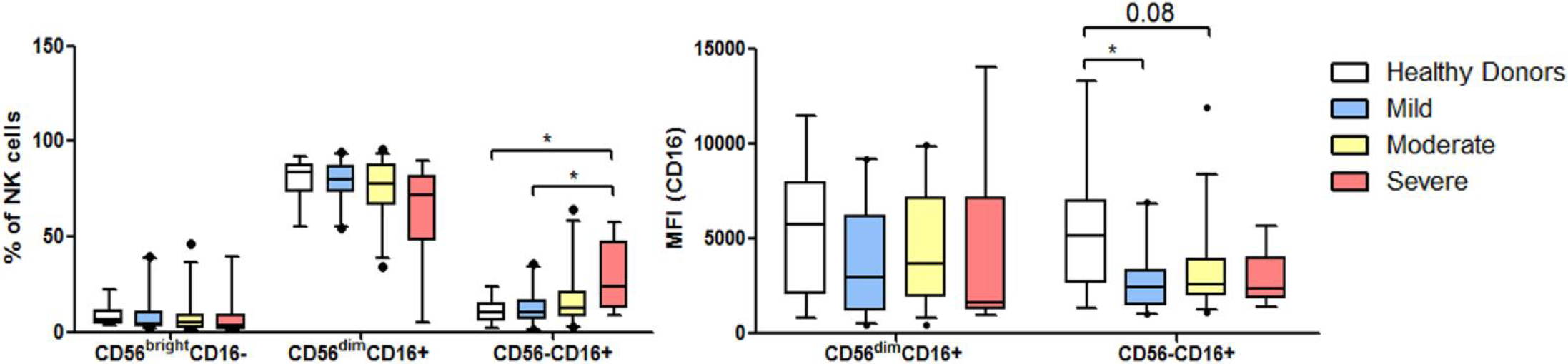
Phenotypic features of NK cells from severe COVID-19 patients. Left: Boxplots show quantification by flow cytometry of the proportion of the three main functional subsets of NK cells in healthy donors (n= 19) and a selected group of COVID-19 patients with different severity degree (mild (n= 29), moderate (n= 40) and severe (n= 15). Right: Boxplot depict CD16 MFI of CD56^dim^CD16+ and CD56-CD16+ NK subpopulations in those same individuals. Asterisks indicate significant differences (p-values for ANOVA Tukey’s contrast test: *p < 0.05, **p < 0.01, ***p < 0.001, **** p < 0.0001)

## Discussion

It has been proposed that the severity of COVID-19 is related to a dysregulation of the immune response to SARS-CoV-2. However, there is a lack of knowledge about the immune profile of COVID-19 patients with different clinical courses. To fill this gap, we have performed a comprehensive characterization of the immune cells populations and soluble mediators of the humoral immunity in the peripheral blood of 276 patients, who presented from mild to critical illness in a single centre during the peak of the pandemic in Madrid, Spain. Plasma cells were significantly elevated in peripheral blood of most COVID-19 patients at the time of admission. In contrast, Ig levels were similar to those of healthy donors, or even decreased in the case of IgG in patients critically ill. Since patients were studied at admission, it was feasible that Ig production had not peaked yet. However, IgG levels decreased further 10 days after admission in patients with longer hospital stays, which suggests that IgG decrease was rather related to severe disease progression. Three possible explanations may account for these apparently contradictory observations. The first explanation is a primary antibody immunodeficiency that debuts with COVID-19. This is supported by the fact that several patients had very low levels of IgG, IgA and IgM in serum both at admission and after 10 days. We have not addressed this possibility yet, however we plan to carry out studies aimed at verifying this hypothesis by assessing Ig levels three months after discharge. Second, it is possible that certain individuals show impaired specific Ig production after infection secondary to generalized lymphopenia, altered B cell maturation, or patient age. Regarding impaired specific Ig production, it is known that sepsis triggers immune cell hyperactivity followed, days later, by immune paralysis, which associates with a poor patient outcome(16). Different mechanisms may underlie this process, including dysregulation of T cell subsets(17). Isotype switching, which results in the production of IgA and IgG antibodies, is a feature of germinal centres in lymphoid follicles following the activation and interaction of specific B cells and Tfh cells by the antigen. Despite the increase in cTfh cells in COVID-19 patients, cTfh from individuals with moderate to severe disease express higher levels of CCR7, which inversely correlates with Tfh capacity for activating B cells(18) as CCR7 prompts their retention in the T zone of the lymph node, preventing their interaction with B cells in the follicle(19). Accordingly, plasma cells significantly decreased after 10 days in patients with prolonged hospitalization. Further experiments to analyze Tfh functional phenotype are needed to confirm a Tfh–dependent defective B cell activation in COVID-19, and their role in a possible immune paralysis at severe stages of the disease.

Likewise, plasma cell number in peripheral blood decreases with age in healthy individuals(20) (unpublished data), which could result in a reduced production of specific neutralizing antibodies and contribute to the severe course in the elderly. Finally, the decrease in circulating IgG could be secondary to its deposit in tissues where they might activate antibody-dependent phagocytosis (ADP), ADCC by NK cells or classical complement pathway. Results from our group(21) demonstrate a profound redistribution of inflammatory monocytes from peripheral blood to the lung where they are probably mediating ADP and tissue injury. Regarding NK cells, several authors have studied the different lymphoid subsets in COVID-19 patients(22-25) and found no differences in the proportion of whole NK cells. However, we observed that the CD56-CD16+ NK cells subset, which has been reported to be particularly efficient at mediating ADCC(15, 26), is greatly expanded in severe patients and could be therefore related to the IgG consumption. Finally, a clear C4 decrease paralleled that of IgG in the same group of patients, thus suggesting a continuous or excessive activation of the classical pathway of complement through specific IgG recognizing viral antigens. The nature of specific antibodies responsible for the activation of complement requires further research. In this regard, a recent study in a SARS-CoV macaque model that appeared before the emergence of first cases of COVID-19, demonstrated that a faster development of neutralizing IgG against SARS-CoV spike characterized animals who developed severe lung injury(12). Moreover, S glycoprotein-specific Ab responses were higher and peaked earlier in deceased patients during the first 15 days after the appearance of symptoms, but dramatically dropped 5 days later, coinciding with clinical deterioration(27). Such timing is similar to that observed in patients with longer hospitalization periods in our study. C4 consumption may be therefore related to the activation of the classical pathway through specific anti-viral antibodies or immune-complexes, or to the activation of the lectins pathway. The absence of significant levels of immune-complexes in sera of these patients does not rule out their local formation and deposit in the tissues. Interestingly, evidence from autopsies and/or biopsies of COVID-19 patients suggests that a widespread complement activation could mediate microvascular injury and leukocyte infiltration, since deposits of different components of the complement system like C5b-9, C4d and MASP are observed in the microvasculature of the lung or the skin(5, 11).

Quantification of C1q and mannose-binding lectin could further clarify which pathways are responsible for C4 cleavage. Whatever the mechanisms of C4 consumption are, our findings support the idea of complement being involved in the hyperinflammatory syndrome observed in COVID-19 patients. In this regard, C5a, one of the main products of the complement cascade activation, is a main proinflammatory molecule, since it has chemoattractant activity, activates most leukocytes and importantly triggers the coagulation cascade. In particular, C5a promotes procoagulant activity through several mechanisms, including the induction of tissue factor by endothelial cells and neutrophils, as well as the upregulation of plasminogen activator inhibitor-1 in mast cells. In addition, the cytolytically inactive terminal complement complex C5b-9 induces procoagulant activity through platelet prothrombinase and activates endothelial cells to express adhesion molecules and tissue factor(28-31).

We find our data of particular therapeutic relevance, since different complement inhibitors are currently being used in the clinics, particularly anti-C5 monoclonal antibody, which has been successfully used in few COVID-19 patients(32, 33).

## Patients and methods

### Study design and population

This is a retrospective observational study including 276 consecutive patients with confirmed detection of SARS-CoV-2 RNA, and admitted to the Accident and Emergency Department of the Hospital Universitario La Princesa because of mild to critical COVID-19 symptoms, from February 27^th^ to April 29^th^.

### Data collection

Demographic and laboratory data described in Table 1 were collected from electronic clinical records and included in an anonymized database. Baseline evaluation of immunity was performed around the 3^rd^ day of admission (median=3 days; percentile 25-75 [p25-p75] 2 to 6). A second evaluation was obtained around the 14th day of admission (median=14 days; p25-p75 12 to 16.5) in a selected group of 37 patients.

### SARS-CoV-2 RNA detection

Samples from nasopharyngeal and throat exudates were obtained with specific swabs as previously described(34). As first line screening, we performed real-time RT-PCR assay targeting the E gene of SARS-CoV-2, with Real Time ready RNA Virus Master on Applied Biosystems TM Quant Studio-5 Real-Time PCR System. This assay was followed by confirmatory testing with the assay TaqPath™COVID-19 CE-IVD Kit RT-PCR Applied Biosystems™(ThermoFisher Scientific, Waltham, MA USA), which contains a set of TaqMan RT PCR assays for *in vitro* diagnostic use. This kit includes three assays that target SARS-CoV-2 genes (Orf1ab, S gene, N gene) and one positive control assay that targets the human RNase P RPPH1 gene(35). Determinations were carried out in an Applied Biosystems™ QuantStudio-5 Real-Time PCR System (CA, USA).

### Sample collection and flow cytometry analysis

Peripheral blood samples from 276 COVID-19 patients were obtained in EDTA tubes for flow cytometry assessment around the 3^rd^ day of admission. The distribution of the different leukocyte subsets was characterized by multiparametric flow cytometry within 24 h after extraction. To this end, 200 µL of whole fresh blood were stained in a BD OneFlow™LST from BD Biosciences (San José, CA, USA) designed to identify and quantify the main lymphocyte populations of T (CD3+, CD4+, CD8+, γδ+), B, and NK lineages. A subgroup of 84 patients were randomly selected during the period of study for an extensive characterization of their lymphocyte subsets. These patients, as well as 19 healthy donors, were studied with an extended panel of multicolor monoclonal antibodies (mAbs): anti-CD4 FITC, anti-CD107 FITC, anti-IgD FITC, anti-PD-1 PE, anti-CD56 PE, anti-CD27 PE, anti-CD38 PerCP/Cy5.5, anti-CXCR5 AF647, anti-IgM APC, anti-CD19 PE-Cy7, anti-CD8 APC/H7, anti-CD10 APC/H7, anti-CCR7 BV421, anti-CD16 V450, anti-CD3 V500 from BD Biosciences (USA); and anti-CD20 V450 from Immunostep (Spain).

Samples were incubated for 30 minutes at room temperature with the respective mAbs. Then, erythrocytes were lysed with FACS lysis solution (BD Biosciences) for 10 minutes and after washing with phosphate buffered saline, samples were analyzed in a FACS Canto II device (BD). At least 100 cells of the less represented subsets were collected and data were analyzed with the FACSDiva and FlowJo Softwares from BD Biosciences. The different leukocyte subsets were assigned according to the strategy showed in supplementary Figure 1.

Data are expressed as percentage or absolute number in cells/ul of a given lymphocyte subpopulation. A dual-platform method was used to calculate absolute cell counts. Mean fluorescence intensity (MFI) was employed to compare the level of expression of certain molecules.

### Serum concentrations of immunoglobulins and complement proteins

Serum samples were tested for total IgG, IgA, IgM, C3 and C4 concentrations by immunonephelometry (Immage800, Beckman Coulter, California, USA). In addition, 19 serum samples from healthy donors obtained during the same period time were used to determine the variability of Ig and C3 and C4 levels in normal conditions. The absence of SARS-Cov-2 infection in healthy donors at the time of extraction was confirmed by retrospective determination of specific antibodies. The presence of circulating immunocomplexes was assessed with C1q CIC ELISA Kit (INOVA, San Diego, USA) following manufacturer’s instructions.

### Statistics

Descriptive results were expressed as mean ± standard deviation (SD) or median and percentile 25-percentile 75 (p25-p75), as appropriate, while qualitative variables are presented as frequency (n) and relative percentages of patients (%). The unpaired, two-tailed, Student t-test was used to compare two independent groups and the paired Student t-test, to analyse two related samples. One-way ANOVA was employed to compare more than two groups and post-hoc multiple comparisons were made with Tukey’s test. Spearman bivariate correlations were performed between serological quantitative markers and cell populations and corrplot R package (available from: https://github.com/taiyun/corrplot) was used for correlation map graphics. Variables in correlation map were reordered using hierarchical cluster method. The p-values were two-sided and statistical significance was considered when p < 0.05. To analyse distribution of lymphocyte in COVID-19 patients and healthy donors, an automated clustering and dimensionality reduction was performed using viSNE and FlowSOM tools (Cytobank). Population cells were normalized using log transformation for analysis. Differences in normalized cells between healthy donors and severity groups (adjusted by sex and age) were assessed with a moderated t-test using limma R package(36). Cell populations that showed significant P-value (FDR = 5%) were considered as differentially expressed between groups. Stata v. 12.0 for Windows and R version 3.5.1 were used for analyses and graphics. GraphPad Prism 4 software was also used for graphics. Data is presented making specification for p < 0.05 (*), p < 0.01 (**), p < 0.001 (***) and p < 0.0001 (****).

### Study approval

This study was approved by the local Research Ethics Committee (register number 4070) and it was carried out following the ethical principles established in the Declaration of Helsinki. All included patients were informed about the study and gave an oral informed consent because of COVID-19 emergency as proposed by AEMPS.

## Data Availability

All data are available upon reasonable request to the corresponding author

## Author contributions

A.A, F.S.M and C.M.C developed the research idea and study concept, designed the study and wrote the manuscript; A.M.J, S.S.A and A.A.S designed and conducted most experiments, analysed the data and prepared the figures. These authors equally contributed to the study; M.S.N performed statistical analysis; M.L.T, E.M.G, M.C, S.C., H. F. and I.G.A. provided clinical data and peripheral blood samples from the study patient cohorts and critically revised the manuscript. All other authors participated in patient samples processing and clinical data collection.

## Acknowledgements

The study was funded by grants SAF2017-82886-R to FS-M from the Ministerio de Economía y Competitividad, and from “La Caixa Banking Foundation” (HR17-00016) to FS-M. Grant PI018/01163 to CMC and grant PI19/00549 to AA were funded by Fondo de Investigaciones Sanitarias, Ministerio de Sanidad y Consumo, Spain. SAF2017-82886-R, PI018/01163 and PI19/00549 grants were also co-funded by European Regional Development Fund, ERDF/FEDER. We thank Dr. Miguel Vicente-Manzanares for proofreading and English editing of the manuscript.

We also thank the immunology service staff for technical support: Victor López-Huete, Alicia Román, Reyes Lázaro-Tejedor, Alicia Vara-Vega, Montserrat Arroyo-Correa, Manuela Mayo.

## References

1. Ruan, Q., Yang, K., Wang, W., Jiang, L., and Song, J. Clinical predictors of mortality due to COVID-19 based on an analysis of data of 150 patients from Wuhan, China: Intensive Care Med. 2020 May;46(5):846–848. doi: 10.1007/s00134-020-05991-x. Epub 2020 Mar 3.

2. Perlman, S., and Dandekar, A.A. Immunopathogenesis of coronavirus infections: implications for SARS. Nat Rev Immunol. 2005 Dec;5(12):917–27. doi: 10.1038/nri1732.

3. Stoermer, K.A., and Morrison, T.E. Complement and viral pathogenesis. Virology. 2011 Mar 15;411(2):362–73. doi: 10.1016/j.virol.2010.12.045. Epub 2011 Feb 2.

4. DÃ¶rner, T., and Radbruch, A. Antibodies and B cell memory in viral immunity. Immunity. 2007 Sep;27(3):384–92. doi: 10.1016/j.immuni.2007.09.002.

5. Gao, T., Hu, M., Zhang, X., Li, H., Zhu, L., Liu, H., Dong, Q., Zhang, Z., Wang, Z., Hu, Y., et al. Highly pathogenic coronavirus N protein aggravates lung injury by MASP-2-mediated complement overactivation. medRxiv:2020.2003.2029.20041962.

6. Nimmerjahn, F., and Ravetch, J.V. Fcgamma receptors as regulators of immune responses. Nat Rev Immunol. 2008 Jan;8(1):34–47. doi: 10.1038/nri2206.

7. Matthay, M.A., Zemans, R.L., Zimmerman, G.A., Arabi, Y.M., Beitler, J.R., Mercat, A., Herridge, M., Randolph, A.G., and Calfee, C.S. Acute respiratory distress syndrome. Nat Rev Dis Primers. 2019 Mar 14;5(1):18. doi: 10.1038/s41572-019-0069-0.

8. Chalmers, S., Khawaja, A., Wieruszewski, P.M., Gajic, O., and Odeyemi, Y. Diagnosis and treatment of acute pulmonary inflammation in critically ill patients: The role of inflammatory biomarkers. World J Crit Care Med. 2019 Sep 11;8(5):59–71. doi: 10.5492/wjccm.v8.i5.59. eCollection 2019 Sep 11.

9. Moore, J.B., and June, C.H. Cytokine release syndrome in severe COVID-19. Science. 2020 May 1;368(6490):473–474. doi: 10.1126/science.abb8925. Epub 2020 Apr 17.

10. Matthay, M.A., and Zemans, R.L. The acute respiratory distress syndrome: pathogenesis and treatment. Annu Rev Pathol. 2011;6:147–63. doi: 10.1146/annurev-pathol-011110-130158.

11. Magro, C., Mulvey, J.J., Berlin, D., Nuovo, G., Salvatore, S., Harp, J., Baxter-Stoltzfus, A., and Laurence, J. Complement associated microvascular injury and thrombosis in the pathogenesis of severe COVID-19 infection: a report of five cases. Transl Res. 2020 Apr 15:S1931-5244(20)30070-0. doi: 10.1016/j.trsl.2020.04.007.

12. Liu, L., Wei, Q., Lin, Q., Fang, J., Wang, H., Kwok, H., Tang, H., Nishiura, K., Peng, J., Tan, Z., et al. Antispike IgG causes severe acute lung injury by skewing macrophage responses during acute SARS-CoV infection. JCI Insight. 2019 Feb 21;4(4):e123158. doi: 10.1172/jci.insight.123158. eCollection 2019 Feb 21.

13. Wu, Z., and McGoogan, J.M. Characteristics of and Important Lessons From the Coronavirus Disease 2019 (COVID-19) Outbreak in China: Summary of a Report of 72â€—314 Cases From the Chinese Center for Disease Control and Prevention. JAMA. 2020 Feb 24. doi: 10.1001/jama.2020.2648.

14. Schmitt, N., and Ueno, H. Blood Tfh cells come with colors. Immunity. 2013 Oct 17;39(4):629–30. doi: 10.1016/j.immuni.2013.09.011.

15. Goodier, M.R., Lusa, C., Sherratt, S., Rodriguez-Galan, A., Behrens, R., and Riley, E.M. Sustained Immune Complex-Mediated Reduction in CD16 Expression after Vaccination Regulates NK Cell Function. Front Immunol. 2016 Sep 26;7:384. doi: 10.3389/fimmu.2016.00384. eCollection 2016.

16. Cheng, S.C., Scicluna, B.P., Arts, R.J., Gresnigt, M.S., Lachmandas, E., Giamarellos-Bourboulis, E.J., Kox, M., Manjeri, G.R., Wagenaars, J.A., Cremer, O.L., et al. Broad defects in the energy metabolism of leukocytes underlie immunoparalysis in sepsis. Nat Immunol. 2016 Apr;17(4):406–13. doi: 10.1038/ni.3398. Epub 2016 Mar 7.

17. Delano, M.J., and Ward, P.A. Sepsis-induced immune dysfunction: can immune therapies reduce mortality? J Clin Invest. 2016 Jan;126(1):23–31. doi: 10.1172/JCI82224. Epub 2016 Jan 4.

18. Schmitt, N., Bentebibel, S.E., and Ueno, H. Phenotype and functions of memory Tfh cells in human blood. Trends Immunol. 2014 Sep;35(9):436–42. doi: 10.1016/j.it.2014.06.002. Epub 2014 Jul 3.

19. Haynes, N.M., Allen, C.D., Lesley, R., Ansel, K.M., Killeen, N., and Cyster, J.G. Role of CXCR5 and CCR7 in follicular Th cell positioning and appearance of a programmed cell death gene-1high germinal center-associated subpopulation. J Immunol. 2007 Oct 15;179(8):5099–108. doi: 10.4049/jimmunol.179.8.5099.

20. Blanco, E., PÃ©rez-AndrÃ©s, M., Arriba-MÃ©ndez, S., Contreras-Sanfeliciano, T., Criado, I., Pelak, O., Serra-Caetano, A., Romero, A., Puig, N., Remesal, A., et al. Age-associated distribution of normal B-cell and plasma cell subsets in peripheral blood. J Allergy Clin Immunol. 2018 Jun;141(6):2208-2219.e16. doi: 10.1016/j.jaci.2018.02.017. Epub 2018 Mar 2.

21. Sanchez-Cerrillo, I., Landete, P., Aldave, B., Sanchez-Alonso, S., Sanchez-Azofra, A., Marcos-Jimenez, A., Avalos, E., Alcaraz-Serna, A., de los Santos, I., Mateu-Albero, T., et al. Differential Redistribution of Activated Monocyte and Dendritic Cell Subsets to the Lung Associates with Severity of COVID-19. medRxiv:2020.2005.2013.20100925.

22. Sun, D.W., Zhang, D., Tian, R.H., Li, Y., Wang, Y.S., Cao, J., Tang, Y., Zhang, N., Zan, T., Gao, L., et al. The underlying changes and predicting role of peripheral blood inflammatory cells in severe COVID-19 patients: A sentinel? Clin Chim Acta. 2020 May 14;508:122–129. doi: 10.1016/j.cca.2020.05.027.

23. Bordoni, V., Sacchi, A., Cimini, E., Notari, S., Grassi, G., Tartaglia, E., Casetti, R., Giancola, L., Bevilacqua, N., Maeurer, M., et al. An inflammatory profile correlates with decreased frequency of cytotoxic cells in COVID-19. Clin Infect Dis. 2020 May 15:ciaa577. doi: 10.1093/cid/ciaa577.

24. Jiang, M., Guo, Y., Luo, Q., Huang, Z., Zhao, R., Liu, S., Le, A., Li, J., and Wan, L. T cell subset counts in peripheral blood can be used as discriminatory biomarkers for diagnosis and severity prediction of COVID-19. J Infect Dis. 2020 May 7:jiaa252. doi: 10.1093/infdis/jiaa252.

25. Giamarellos-Bourboulis, E.J., Netea, M.G., Rovina, N., Akinosoglou, K., Antoniadou, A., Antonakos, N., Damoraki, G., Gkavogianni, T., Adami, M.E., Katsaounou, P., et al. Complex Immune Dysregulation in COVID-19 Patients with Severe Respiratory Failure. Cell Host Microbe. 2020 Apr 17:S1931-3128(20)30236-5. doi: 10.1016/j.chom.2020.04.009.

26. Costa-Garcia, M., Vera, A., Moraru, M., Vilches, C., LÃ^3^pez-Botet, M., and Muntasell, A. Antibody-mediated response of NKG2Cbright NK cells against human cytomegalovirus. J Immunol. 2015 Mar 15;194(6):2715–24. doi: 10.4049/jimmunol.1402281. Epub 2015 Feb 9.

27. Zhang, L., Zhang, F., Yu, W., He, T., Yu, J., Yi, C.E., Ba, L., Li, W., Farzan, M., Chen, Z., et al. Antibody responses against SARS coronavirus are correlated with disease outcome of infected individuals. J Med Virol. 2006 Jan;78(1):1–8. doi: 10.1002/jmv.20499.

28. Wojta, J., Kaun, C., Zorn, G., Ghannadan, M., Hauswirth, A.W., Sperr, W.R., Fritsch, G., Printz, D., Binder, B.R., Schatzl, G., et al. C5a stimulates production of plasminogen activator inhibitor-1 in human mast cells and basophils. Blood. 2002 Jul 15;100(2):517–23. doi: 10.1182/blood.v100.2.517.

29. Wiedmer, T., Esmon, C., and Sims, P. 1986. On the mechanism by which complement proteins C5b-9 increase platelet prothrombinase activity. The Journal of biological chemistry 261:14587–14592.

30. Ikeda, K., Nagasawa, K., Horiuchi, T., Tsuru, T., Nishizaka, H., and Niho, Y. C5a induces tissue factor activity on endothelial cells. Thromb Haemost. 1997 Feb;77(2):394–8.

31. Tedesco, F., Pausa, M., Nardon, E., Introna, M., Mantovani, A., and Dobrina, A. The cytolytically inactive terminal complement complex activates endothelial cells to express adhesion molecules and tissue factor procoagulant activity. J Exp Med. 1997 May 5;185(9):1619–27. doi: 10.1084/jem.185.9.1619.

32. Diurno, F., Numis, F.G., Porta, G., Cirillo, F., Maddaluno, S., Ragozzino, A., De Negri, P., Di Gennaro, C., Pagano, A., Allegorico, E., et al. Eculizumab treatment in patients with COVID-19: preliminary results from real life ASL Napoli 2 Nord experience. Eur Rev Med Pharmacol Sci. 2020 Apr;24(7):4040–4047. doi: 10.26355/eurrev_202004_20875.

33. Mastaglio, S., Ruggeri, A., Risitano, A.M., Angelillo, P., Yancopoulou, D., Mastellos, D.C., Huber-Lang, M., Piemontese, S., Assanelli, A., Garlanda, C., et al. The first case of COVID-19 treated with the complement C3 inhibitor AMY-101. Clin Immunol. 2020 Apr 29;215:108450. doi: 10.1016/j.clim.2020.108450.

34. Wang, W., Xu, Y., Gao, R., Lu, R., Han, K., Wu, G., and Tan, W. Detection of SARS-CoV-2 in Different Types of Clinical Specimens. JAMA. 2020 Mar 11;323(18):1843–4. doi: 10.1001/jama.2020.3786.

35. Lu, R., Zhao, X., Li, J., Niu, P., Yang, B., Wu, H., Wang, W., Song, H., Huang, B., Zhu, N., et al. Genomic characterisation and epidemiology of 2019 novel coronavirus: implications for virus origins and receptor binding. Lancet. 2020 Feb 22;395(10224):565–574. doi: 10.1016/S0140-6736(20)30251-8. Epub 2020 Jan 30.

36. Ritchie, M.E., Phipson, B., Wu, D., Hu, Y., Law, C.W., Shi, W., and Smyth, G.K. limma powers differential expression analyses for RNA-sequencing and microarray studies. Nucleic Acids Res. 2015 Apr 20;43(7):e47. doi: 10.1093/nar/gkv007. Epub 2015 Jan 20.

